# Polygenic Risk Scores for Alcohol Involvement Relate to Brain Structure in Substance-Naïve Children: Results from the ABCD Study

**DOI:** 10.1101/2020.07.25.20162032

**Authors:** Alexander S. Hatoum, Emma C. Johnson, David A.A. Baranger, Sarah E. Paul, Arpana Agrawal, Ryan Bogdan

**Affiliations:** Washington University St. Louis Medical School, Department of Psychiatry, Pittsburgh, PA; Washington University St. Louis, Department of Psychology & Brain Sciences, Pittsburgh, PA; Department of Psychiatry, University of Pittsburgh Medical Center, Pittsburgh, PA

## Abstract

In substance naïve children (n=3,013), polygenic risk score (PRS) for problematic alcohol use was associated with lower volume of the frontal pole and greater cortical thickness of the supramarginal gyrus. Several other areas showed nominal significance. These associations suggest that genetic liability to alcohol involvement may manifest as variability in brain structure prior to consumption of the first alcoholic drink and alter brain morphometry during the start of adolescence.

---

Excessive alcohol use is an escalating international health problem that accounts for over 5% of global deaths and disease burden (World Health Organization, 2018). Alcohol use and use disorders have been reliably associated with magnetic resonance imaging (MRI)-derived brain structure phenotypes, particularly among regions and pathways that feature prominently in executive function, incentive salience, and negative emotionality (Hampton et al., 2019; Mackey et al., 2019). Consistent with evidence from non-human animal models, it is widely speculated that these structural reductions arise as a consequence of chronic alcohol exposure and contribute to the development of alcohol-related comorbidities (e.g., risk taking, Alzheimer’s Disease)(Crews, 2008; Yu et al., 2016). However, emerging data challenging this common interpretation suggest that these brain structure correlates may, at least partially, reflect genetically conferred predisposing risk factors for alcohol involvement (Baranger et al., 2019; Robert et al., 2019).

Building upon twin and family studies documenting the moderate heritability of alcohol use and use disorders (30-50%) (Verhulst et al., 2015; Walters, 2002), large-scale genome-wide association studies (GWASs) have begun to reliably characterize their polygenic architecture. Polygenic risk scores (PRS) that effectively capture this polygenic liability can be combined with neuroimaging data to investigate whether individual differences in neural phenotypes may be partially attributable to common underlying genomic vulnerability and/or arise following substance exposure, use, and/or problematic use. However, given the ubiquity of lifetime alcohol use, efforts to disentangle such predispositional effects from neurotoxic consequences of chronic exposure have been limited.

Here, among 3,013 substance-naïve^a^ children (age=9.92±0.62 years, 47% Female; 100% genomically-confirmed European ancestry; **Supplemental Table 1**) who completed the baseline session of the ongoing Adolescent Brain Cognitive Development (ABCD) study (data release 2.0.1), we test whether PRS for typical alcohol use and problematic use are associated with brain structure phenotypes. To this end, we generated PRS using the largest available GWASs of alcohol use (i.e., alcohol drinks/week [DPW-PRS], training n=537,349) and problematic use (i.e., alcohol use disorder/problem use [PAU-PRS], training n=352,365) and considered MRI-derived brain structure phenotypes that have been previously linked to substance involvement and related phenotypes (i.e., total and regional: cortical thickness, surface area and volume; subcortical volume; white matter volume and integrity). Mixed effects regressions were nested by family and scanner and included the following fixed covariates in all analyses: age, sex, age by sex, first 20 ancestrally-informative PCs, genotyping batch, socioeconomic status proxies (caregiver education, combined family income, and parent marital status were all considered as separate covariates), and prenatal substance exposure. False-discovery rate (FDR) correction within modality (e.g., cortical thickness) was used to adjust for multiple testing. We hypothesized that PRS for typical and problematic alcohol use would be associated with brain structure metrics previously linked to alcohol involvement, even in the absence of alcohol exposure. Further, we expected that variability in brain structure would indirectly link alcohol involvement PRS to related behavioral risk factors (i.e., cognitive performance, internalizing symptoms, externalizing symptoms). Methodological details are provided in **Online Methods**. Results surviving FDR correction are discussed here; all results are presented in the **Supplement**.

Consistent with evidence of lower frontal pole gray matter volume among adolescents who drink heavily(Heikkinen et al., 2017), *PAU-PRS* were associated with reduced frontal pole gray matter volume (standardized β= -0.054, r^2^=0.003, p_fdr_=0.031; **Figure 1**; **Table 1**). Notably, DPW-PRS showed a similar pattern of findings, albeit at nominal levels of significance (**Table 1**); given that typical and problematic alcohol use show similar neural correlates(Paul et al., 2008) and are genetically correlated (rg=0.78),(Zhou et al., 2020) such consistency is unsurprising. Results from a regression with PAU-PRS and DPW-PRS entered simultaneously show that this finding is driven primarily by PAU-PRS (**Online Methods; Supplementary Table 1**). However, unlike other studies showing reduced volume is associated with increased (Gröpper et al., 2016) externalizing behavior, we found no evidence that externalizing or other putative alcohol-related behaviors (e.g., cognitive performance) were associated with frontal pole volume.

**Figure 1.**
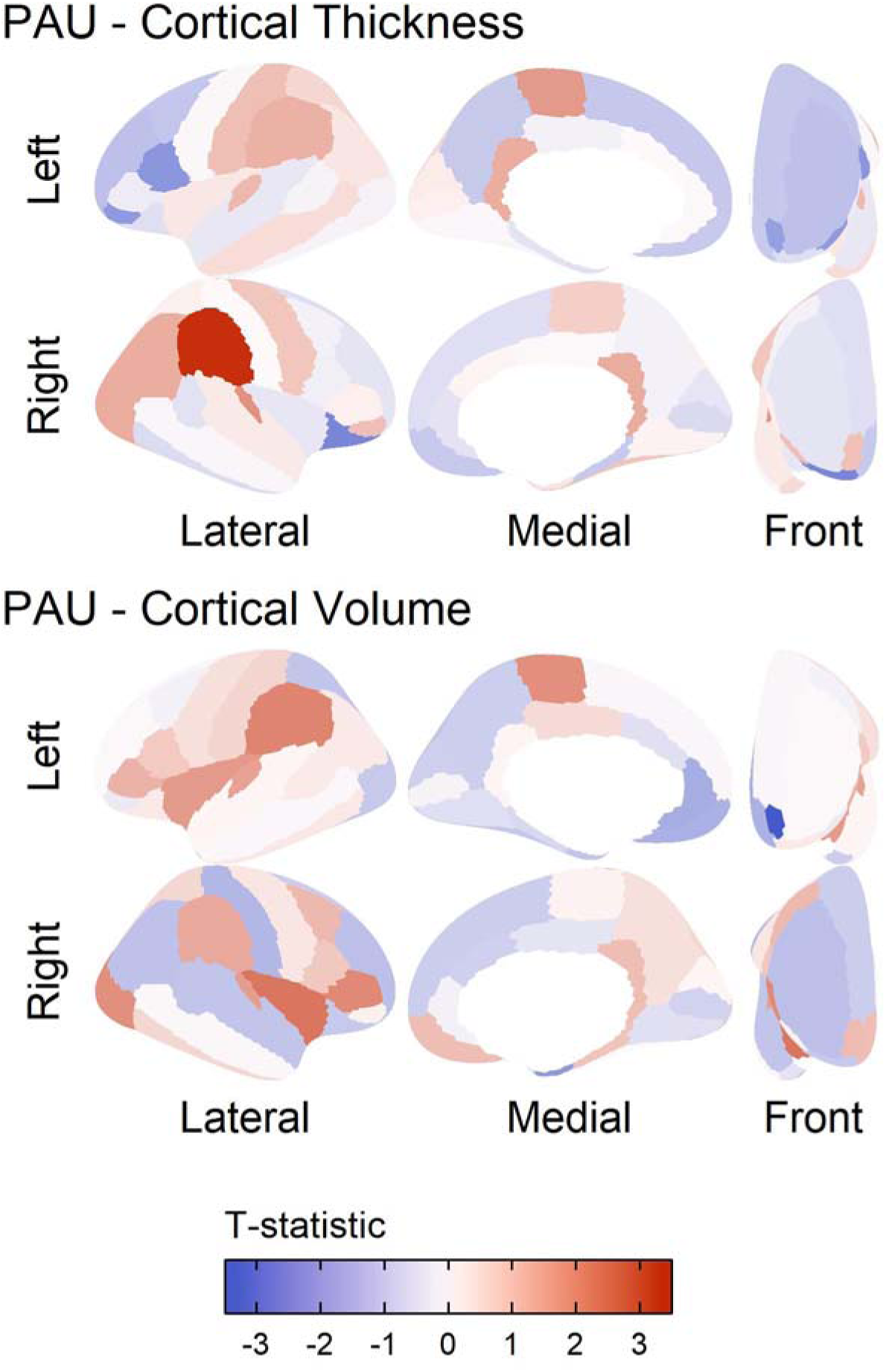
Results of polygenic risk prediction for Problematic Alcohol Use (PAU) plotted on the cortex. The results are shown for those modalities that had significant regions in the Desikan atlas after controlling for mean of the modality, prenatal alcohol exposure, age, sex, age by sex, first 20 principal components, SES, and accounting for family and site as random nested effects. Red indicates areas where higher PAU polygenic risk predicted more of that modality. In the top panel, we focus on supramarginal gyrus cortical thickness prediction by PRS, as this association was significant after multiple corrections. In the bottom panel, we focus on volume of the temporal pole, as this was significantly associated with PAU PRS after multiple correction.

**Table 1.** Nominally significant results for the PAU and DPW PRS Prediction

| **PAU** |  |  |  |  |
| --- | --- | --- | --- | --- |
| Modality | Region | Beta | STE | Nominal P-value |
| Area | Right Insula | 0.027 | 0.014 | 0.046 |
| Area | Right Pars Triangularis | 0.035 | 0.016 | 0.033 |
| Thickness | Left Pars Orbitalis | -0.031 | 0.016 | 0.049 |
| Thickness | Left Pars Opercularis | -0.028 | 0.014 | 0.043 |
| Thickness | Right Lateral Orbital Frontal Gyrus | -0.035 | 0.015 | 0.017 |
| Thickness | Right Supramarginal Gyrus | 0.032 | 0.010 | 0.001* |
| Volume | Left Frontal Pole | -0.057 | 0.017 | 0.001* |
| fractional anisotropy | right anterior thalamic radiations | -0.024 | 0.012 | 0.046 |
| fractional anisotropy | left anterior thalamic radiations | -0.025 | 0.012 | 0.030 |
| White Matter volume | Corpus Callosum | -0.010 | 0.005 | 0.045 |
| White Matter volume | Right Parahippocampal cingulum | -0.030 | 0.014 | 0.041 |
| White Matter volume | Forceps Minor | -0.023 | 0.009 | 0.014 |
| **DPW** |  |  |  |  |
| Area | Right Insula | 0.031 | 0.014 | 0.023 |
| Area | Left Superior Parietal Gyrus | -0.033 | 0.013 | 0.013 |
| Thickness | Right Inferior Parietal Gyrus | 0.024 | 0.010 | 0.012 |
| Thickness | Right Pericalcarine Cortex | -0.034 | 0.016 | 0.039 |
| Thickness | Right Temporal Pole | -0.033 | 0.017 | 0.048 |
| Volume | Right Entorhinal Cortex | -0.047 | 0.018 | 0.008 |
| Volume | Left Insula | 0.029 | 0.013 | 0.024 |
| Volume | Right Insula | 0.027 | 0.013 | 0.039 |
| Volume | Left Superior Parietal Gyrus | -0.029 | 0.014 | 0.044 |
| Volume | Left Frontal Pole | -0.034 | 0.017 | 0.049 |
| Volume | Left Pallidum | -0.030 | 0.015 | 0.049 |
| Mean Diffusivity | Right Parahippocampal Cingulum | -0.033 | 0.016 | 0.041 |
| Mean Diffusivity | Right Inferior Longitudinal Fasciculus | 0.029 | 0.013 | 0.029 |

**Note.** Nominal significant results shown by region across all modalities. STE= Standard Error.
\* = Area was significant after FDR correction.

**Table 2.**
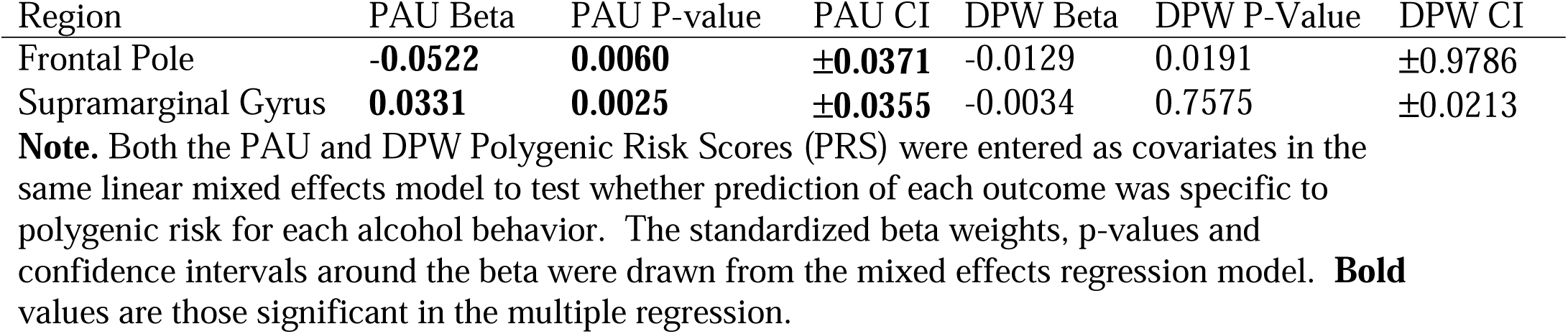
Problematic Alcohol Use (PAU) and Drinks Per Week (DPW) PRS Prediction of Top Outcomes in Multiple Linear Model, Accounting for the Other PRS

| Region | PAU Beta | PAU P-value | PAU CI | DPW Beta | DPW P-Value | DPW CI |
| --- | --- | --- | --- | --- | --- | --- |
| Frontal Pole | **-0.0522** | **0.0060** | **±0.0371** | -0.0129 | 0.0191 | ±0.9786 |
| Supramarginal Gyrus | **0.0331** | **0.0025** | **±0.0355** | -0.0034 | 0.7575 | ±0.0213 |

**Note.** Both the PAU and DPW Polygenic Risk Scores (PRS) were entered as covariates in the same linear mixed effects model to test whether prediction of each outcome was specific to polygenic risk for each alcohol behavior. The standardized beta weights, p-values and confidence intervals around the beta were drawn from the mixed effects regression model. **Bold** values are those significant in the multiple regression.

We also found that PAU-PRS were associated with greater supramarginal gyrus cortical thickness (standardized β=0.009, r^2^=0.001, p_fdr_=0.045; **Figure 1; Table 1**).^b^ Greater supramarginal gyrus cortical thickness has been found in substance-naïve children of alcohol dependent individuals and temporally predicts the initiation of regular alcohol use during adolescence (Holla et al., 2019; Jacobus et al., 2016). In contrast, lower supramarginal gyrus cortical thickness has been observed among individuals with alcohol dependence (Mackey et al., 2019). Together, this evidence raises the intriguing possibility that some brain structure correlates of alcohol involvement may be developmentally dependent. Indeed, throughout childhood and adolescence neuronal pruning appears to support long term planning, working memory performance, language, and attention (Selemon, 2013) and it has been postulated that greater indices of gray matter among those at familial risk for alcohol problems may reflect a developmental delay in neural pruning that places adolescents at risk for substance use, which, in turn, accelerates neuronal pruning in later development (Winters and Arria, 2011). Given that greater cortical thickness of the supramarginal gyrus is present among those at elevated risk for problematic alcohol use and this region supports cognition and language (Stoeckel et al., 2009), this is plausible. However, supramarginal gyrus cortical thickness was not significantly correlated with cognition (**Supplement**); it remains possible that such associations are developmentally dependent and coincide with alcohol experimentation. No brain regions were discovered for typical alcohol use after correction for multiple comparisons (DPW-PRS, supplemental Results).

Several other nominally significant associations arose in analyses (**Table 1**). First, the inferior and superior parietal cortex and the insula have been previously associated with substance dependence in large mega-analyses (Mackey et al., 2019), but in the opposite direction of those discovered. This mirrors our findings with the supramarginal gyrus, as the results show opposite patterns in adults, but replicate past findings in adolescence that find these same directions of effect in likelihood of initiation and in familial risk for AUD.(Holla et al., 2019; Jacobus et al., 2016). It is possible that brain regions exerting the most dominant influence may show fluid changes across development, and that individual difference genetics as well as alcohol exposure(Shnitko et al., 2019) may drive developmentally dependent differences.

Here, among 3,013 substance-naïve children, we find evidence that alcohol-related differences in brain structure may, at least partially, reflect genetically-conferred predipsositional risk factors that precede alcohol exposure. These findings, combined with evidence from humans and non-human animal models challenge widespread speculation that brain-based associations with alcohol are solely attributable to the neurotoxic effects of alcohol (Baranger et al., 2019). It is important to consider the limitations of this study. First, while the study of alcohol and substance naïve children allowed us to preclude that associations may be attributable to alcohol exposure, we were unable to test whether brain structure correlates of genomic risk are associated with onsets of alcohol use and other drinking milestones. As the longitudinal ABCD study continues to collect data, it will be particularly interesting to examine how the interplay between polygenic risk and exposure influence brain structure trajectories, and vice versa (i.e., how malleability of brain development modulates escalation or desistance in alcohol use). Second, PRS typically have low cross-trait (i.e., from GWAS phenotype to a related phenotype) predictive utility. While our findings suggest that variability in brain structure may represent predispositional biomarkers for alcohol involvement, the small effect sizes (i.e., maximum R^2^=0.003) and resource intensive nature of neuroimaging limit their clinical utility. Further, these small effects combined with multiple testing burden may have contributed to false negative, and we should not conclude that a null effect here means an association in adults is due to neurotoxicity. Third, as the discovery GWAS were of individuals of predominantly European descent, we restricted our ABCD sample to children of similar ancestral background. Our analyses in the African-ancestry sample of ABCD revealed no significant results (supplemental table 3), likely due to low power of the discovery GWAS of matched ancestry (N = 62,447 vs. 352,365 for the European-ancestry GWAS). This highlights the great need for more non-European discovery samples.

Limitations notwithstanding, our study provides initial evidence that alcohol-related brain structure correlates represent genomic risk factors for alcohol involvement. This does not preclude the possibility of neurotoxic effects, as we have not tested the reverse. Rather, this bolsters the argument that genetic vulnerability is conveyed via alterations in brain structure and that these alterations are present during middle childhood.

## Data Availability

Data is all fully publicly available as part of the ABCD study data release.

**The Mllion Veterans Project summary statistics were accessed via dbGaP (phs001672**.**v1**.**p1) as part of #24806: Neurobiological bases of psychiatric traits**

**T**he authors thank Million Veteran Program (MVP) staff, researchers, and volunteers, who have contributed to MVP, and especially participants who previously served their country in the military and now generously agreed to enroll in the study. (See https://www.research.va.gov/mvp/ for more details).

The citation for MVP is Gaziano, J.M. et al. Million Veteran Program: A mega-biobank to study genetic influences on health and disease. J Clin Epidemiol 70, 214-23 (2016).

This research is based on data from the Million Veteran Program, Office of Research and Development, Veterans Health Administration, and was supported by the Veterans Administration (VA) Cooperative Studies Program (CSP) award #G002.

ASH receives support from DA007261-17. ECJ receives support from AA027435-02. AA receives support from MH109532 and K02DA032573. DAAB receives support from MH018951. Dr. Bogdan (AG052564, AA027827, DA046224). ASH, ECJ, AA, and RB developed the research questions. ASH conducted analyses, with help from ECJ, DAAB, and SEP. ASH and RB drafted the manuscript. ECJ, AA, DAAB, and SEP provided critical revision of the manuscript for important intellectual content. ASH had full access to all data in the study and take responsibility for the integrity of the data and accuracy of the data analyses. Conflict of interest disclosures: No disclosures were reported.

## Online Methods

### Sample

The Adolescent Brain and Cognitive Development Study (ABCD) is an ongoing multi-site longitudinal study of child health and development. Children (N=11,875), ages 8.9-11 recruited from 22^c^ sites across the nation, completed a baseline assessment of child behavior, health and cognitive assessment. The study included a twin design, in which 2,108 participants were twins and 30 were triplets. For our analyses, the sample was restricted to those of genomically-confirmed European ancestry (n=4,737 using stringent criteria; details below) who self-reported no exposure to substances and screened negative for substances according to hair toxicology, leaving a final analytic sample (n=3,434 individuals, mean age =9.92 years, std age = 0.62, 47 % Female, n=3,013 with no missing data).

### Measures

#### Brain Structure. MRI Acquisition and Processing

Of the 11,875 ABCD participants, 11,556 have structural MRI data. A detailed description of the ABCD Study imaging acquisition protocol and parameters can be found in Casey et al. (2018)(Casey et al., 2018). Briefly, 3 T (Siemens, Phillips and GE) MRI scanners were used to obtain 1 mm isotropic T1-weighted structural, using either a 32-channel head or 64-channel head-and-neck coil. In order to minimize variability across scanners, MRI scan protocols were carefully harmonized across the three MRI vendor platforms. Because head motion is more common in imaging of pediatric populations, real-time motion detection and correction were implemented to mitigate this concern (prospective motion correction on the GE and Volumetric Navigators on the Siemens platforms).

A detailed description of ABCD Study image processing and analysis methodology can be found in Hagler et al., (2019). The Multi-Modal Processing Stream software package, which includes FreeSurfer 5.3, was used to process MRI data. The ABCD processing pipeline specifies a modified intensity normalization process, and the standard FreeSurfer cortical and subcortical reconstruction pipeline was implemented in addition to generate structural measures such as volume and cortical thickness. Hagler et al. (2019) provides a comprehensive description of the quality-control measures conducted on the processed imaging data. Only participants whose structural MRI reconstructions passed these QC tests (n=11,076) were retained. Cortical gray matter volume, thickness, and surface area aligned to the Desikan atlas and white matter volume, mean diffusivity and fractional anisotropy along AtlasTrack were extracted from ABCD data release 2.0.1, which rectified an issue with data release 2.0 with regard to the laterality of data in some participants.

We extracted estimates separately per hemisphere. Further, for cortical gray matter we chose to include cortical thickness, surface area and volume aligned to the Desikan atlas through the Freesurfer protocol because this method better distinguishes white and gray matter than traditional VBM approaches (Fischl et al., 1999). We ran models using the Desikan atlas parcellation for the cortex, within each hemisphere (34 brain regions). For white matter modalities, we used fractional anisotropy and mean diffusivity because they represent the primary components of many different white matter metrics obtain from Diffusion Tensor imaging and they are the most commonly used for imaging research.

##### Genotyping, Quality Control, and Imputation

The Rutgers University Cell and DNA repository genotyped saliva samples on the Smokescreen array (Baurley et al., 2016). Genotyped calls were aligned to GRCh37 (hg19), and all individuals self-reporting ancestral origins (i.e., self-reported race) other than Europeans or African Americans were excluded because these were the only ancestral populations in which GWAS summary statistics are available and evidence that the predictive utility of polygenic risk scores suffers when applied across ancestral origins (Bogdan et al., 2018). we separated individuals into self-reported populations before genotype QC. QC steps also were used to confirm that individuals belonged to these ethnic groups using their genomic data.

The following preprocessing steps were conducted with the Ricopili pipeline (Lam et al., 2019): SNPs with call rates ≥ 0.95 and MAF ≥ 1% were retained. Individuals with high rates of missingness (>5%) and autosomal heterozygosity deviation (F_HET_) outside of ± 2 SD were removed. After sample QC, SNPs were further filtered to call rate ≥ 0.98 and Hardy-Weinberg p-values > 1E-6 (founders only), which yielded 372,342 SNPs. In order to reconcile mismatches, sex checks were conducted with follow-up.

Individuals whose data passed the first phase of QC were then checked for relatedness--both known and cryptic--and Mendelian errors were resolved. Next, using data from unrelated individuals (pi-hat ≤ 0.20) and an LD pruned set of common (MAF>0.05) and non-palindromic SNPs (and excluding MHC and chromosome 8 inversion region), principal components analysis (PCA) was performed in EIGENSTRAT using the European 1000 Genomes Project phase 3 data. Only those individuals whose data aligned with ancestral non-Hispanic European ancestry were retained, yielding a sample of 4,737. Due to the sensitivity of the PRS approach to admixture, we took a conservative approach and performed stringent exclusion for ancestral outliers, consistent with the Psychiatric Genomics Consortium’s Ricopili pipeline (Lam et al., 2019). After selection, a final ancestrally-informative PCA was conducted, and the first 20 PCs were projected from founders to other relatives. Imputation to 1000 Genomes and Haplotype Reference Consortium (HRC) data for Europeans and KAPPA for African Americans was conducted using strictly QCed SNPs on the Michigan Imputation Server, yielding 39,127,678 SNPs. Dosage data were converted to hard-call genotypes using Plink, and only SNPs with imputation r^2^ scores ≥ 0.3 were used to create polygenic scores.

### Polygenic Risk Scores

Polygenic Risk Scores (PRS) were generated using the PRS-cs software package (Ge et al., 2019). The PRS-cs auto approach calculates PRS by assuming a general distribution of effect sizes across the genome, and then reweighting SNPs based off of this assumption, their effect size in the original GWAS, and their linkage disequilibrium (LD) patterns to create weights for every SNP that are then summed for a final score. We chose PRS-cs auto because we wanted to avoid the potential over-fitting to one particular PRS threshold and multiple-testing burdens of P+T, as well as the potential gain of using ALL SNPs, rather than only SNPs below an arbitrary p-value cutoff. Our models were trained using two well-powered GWAS from major consortia, specifically Drinks per week (DPW, original GWAS N=537,349 (Liu et al., 2019)) and a problematic alcohol use (PAU) meta-analysis of GWASs of alcohol use disorder (specifically, alcohol dependence from the Psychiatric Genomics Consortium (Walters et al., 2018) and ICD codes for alcohol use disorder from the Million Veteran Program (Zhou et al., 2020) and a GWAS of scores on the problem subscale of the Alcohol Use Disorders Identification Test (AUDIT-P)(Sanchez-Roige et al., 2019) (Original GWAS N=352,391) from the UK Biobank. We chose to use both DPW-PRS and PAU-PRS because of past work that shows genetic separability in use and use disorder for drinking (Walters et al., 2018).

### Behavioral Measures

For each discovered brain region, we ran a test of the association between that brain region and cognitive ability, as indexed by full-scale IQ, fluid reasoning, crystalized reasoning, as well as internalizing and externalizing symptoms. Measures for IQ were from the NIH Toolbox (Gershon et al., 2013), and internalizing and externalizing scales were taken from the Child Behavior Checklist (CBCL) (Achenbach et al., 1987).

### Statistical Analysis

We associated each PRS with each brain region with a mixed-effects model estimated in the lme4 (Bates et al., 2015) package in R. To account for non-independence of individuals due to clustering in families and to account for scanner-specific effects at each site, we nested data by family and scanner MRI serial number as random effects. We controlled for the first 20 ancestry principal components, mean of each modality (separately when predicting regions of that modality, i.e., mean thickness predicting regional thickness, mean surface area predicting regional surface area), prenatal exposure, age, sex, age by sex, caregiver education, combined caregiver income and batch as fixed effects in each model. We removed genotyping batch 461 as well as 111 individuals who did not pass quality control on recommendation from the ABCD project. We ran a test of association within each modality and separately for each brain region with each PRS. We corrected our results for multiple comparisons by using FDR across all brain measures within a modality (Gray matter cortex = 34 regions, white matter = 36 tracts, subcortex = 26 regions).

## Footnotes

a Children self-reported no substance use (including substances other than alcohol or tobacco) and screened negative according to hair toxicology (see **Supplementary Information for a table of exclusions**). All participants were required to have non-missing data on all variables used in analyses.

b Analyses in individuals of African ancestry (n=898) using PAU GWAS results from an African ancestral discovery (training n= 62,447) yielded no significant associations, presumably due to low power (**Online Methods; Supplemental Table 3**). GWAS summary statistics for Drinks Per Week are not currently available for those of African ancestry.

c Cornell University was an original collection site that collected data from 34 participants, before being moved to Yale University. ABCD documentation reports 21 data collection sites and does not list Cornell; our analyses nested data based on 22 data collection sites, including the original Cornell site.

## Notes

### Competing Interest Statement

The authors have declared no competing interest.

### Author Declarations

Data are publicly available as part of the ABCD study, IRB approval was not needed.

